# Multidimensional symptom burden across sleep, pain, affect, cognition and energy: cross-sectional associations with chronic pain interference and work disability in UK Biobank

**DOI:** 10.64898/2026.08.03.26359604

**Authors:** Sarah Ciechanowicz, Kangning Li, Daqing Ma

**Author notes:** Corresponding author: Sarah Ciechanowicz, Department of Anaesthesia and Perioperative Medicine, University College London Hospital, 235 Euston Road, London NW1 2BU, UK.

## Abstract

**Background:** Sleep, pain, affect, cognition and energy (SPACE) has been described as a latent symptom-severity construct in chronic overlapping pain conditions. Its population-level structure and associations with chronic pain interference and functional disability remain uncertain.

**Methods:** We conducted a cross-sectional analysis of UK Biobank participants aged ≥40 years. Prespecified standardised symptom domains were examined using correlation analysis, principal component analysis, exploratory factor analysis and k-means clustering. Associations with chronic pain interference, current work disability, poor self-rated health and longstanding illness or disability were assessed using covariate-adjusted logistic regression. A four-domain score excluding pain tested whether associations extended beyond the pain domain. Secondary analyses examined convergence with actigraphy, biomarkers, polygenic risk scores and brain magnetic resonance imaging phenotypes.

**Results:** Of 501,935 eligible participants, 475,134 had complete domain data. Domains were modestly intercorrelated (r=0.04–0.46). Clustering identified lower- and higher-burden phenotypes comprising 70.5% and 29.5% of participants; current work disability occurred in 1.3% and 10.3%, respectively. Adding the four non-pain domains increased the area under the curve for chronic pain interference from 0.593 to 0.672 and for work disability from 0.748 to 0.850. Inclusion of pain increased the work-disability area under the curve to 0.863. Multimodal measures added smaller increments.

**Conclusions:** A multidimensional symptom profile was identifiable at population scale and was concurrently associated with chronic pain interference and work disability, with non-pain domains contributing information beyond pain burden alone.

**Significance:** Chronic pain interference and work disability were associated with a broader multidimensional symptom burden spanning sleep, affect, cognition and energy, rather than pain alone. In a large population cohort, the non-pain domains added substantial concurrent discriminatory information. These findings support multidimensional assessment of functionally impairing pain and justify prospective evaluation of whether symptom-system profiles predict incident disability, healthcare use, recovery trajectories or treatment response.

## 1 Introduction

Schrepf and colleagues identified SPACE (Sleep, Pain, Affect, Cognition and Energy) as a latent symptom-severity dimension in chronic overlapping pain conditions (Schrepf et al., 2018). In the Multidisciplinary Approach to the Study of Chronic Pelvic Pain Research Network, SPACE emerged alongside a distinct generalised sensory-sensitivity dimension and showed consistency across cohorts and longitudinal assessments, supporting the interpretation of constitutional symptom burden as one dimension of centralised pain presentations (Schrepf et al., 2018).

Subsequent studies have extended this work. In UK Biobank participants without recent or chronic pain at baseline, a greater number of central nervous system-associated symptoms was associated with a higher subsequent incidence of chronic primary pain, suggesting that multisymptom burden may precede the development of some chronic pain conditions (Kelleher et al., 2025b). A population-level neuroimaging study subsequently examined SPACE symptoms as potential mediators of associations between descending pain modulatory system connectivity and Fibromyalgia Index scores, which were used as a dimensional marker of nociplastic pain severity (Kelleher et al., 2026). Nociplastic pain severity has also been associated cross-sectionally with poorer executive function (Kelleher et al., 2025a). Latent-class analysis applying the SPACE domains in premenarchal females further suggests that the framework may be relevant across developmental contexts (Smith et al., 2025).

Perioperative data provide preliminary translational support. In a single-centre prospective cohort following caesarean delivery, a five-domain SPACE model was associated with chronic pain at 3 months, suggesting that early multidomain symptom burden may be relevant to persistent postsurgical pain (Ciechanowicz et al., 2026).

However, population-level evaluation of SPACE as an integrated five-domain construct remains limited. Existing UK Biobank studies have examined counts of related central nervous system-associated symptoms or modelled individual SPACE symptoms in relation to chronic pain and brain connectivity, rather than formally evaluating the structural coherence of an operationalised five-domain construct across the broader adult cohort (Kelleher et al., 2025b; Kelleher et al., 2026). Whether multidomain SPACE burden is associated with work disability and other functional outcomes, and whether it shows consistent associations across multiple external measurement modalities, therefore remains uncertain.

Using UK Biobank, we evaluated SPACE as a population-level transdiagnostic construct. We examined the structural coherence of five operationalised SPACE domains, assessed associations between SPACE burden and functional impairment, and explored convergence with multimodal markers. We hypothesised that SPACE would show evidence of a shared multidimensional structure, would be associated with functional impairment, and would demonstrate directionally consistent associations with measures of physical activity, neurobiology, genetic liability and physiology.

## 2 Methods

### 2.1 Study design and population

This was a population-based, cross-sectional observational study using UK Biobank baseline assessment data (baseline recruitment 2006–2010), with linked health-record data, follow-up questionnaire data, genetic data, imaging-derived phenotypes, wrist-worn accelerometry, and blood and physiological biomarkers used in secondary analyses where available. For the primary analyses, the SPACE symptom domains and the clinical evaluation outcomes were ascertained cross-sectionally from the same baseline assessment, and associations are therefore concurrent rather than predictive. The primary aim was to evaluate whether an analogous SPACE symptom construct may be operationalised at population scale and whether it demonstrated evidence of structural coherence, clinical associations and exploratory multimodal convergence. The study is reported in accordance with the STROBE guideline for cross-sectional studies. Model-discrimination statistics were used to quantify the concurrent, incremental information contributed by SPACE.

The primary analytic population comprised UK Biobank participants aged ≥40 years with available baseline assessment-centre data. Participants contributed to structural analyses if complete data were available for the baseline SPACE domains. Complete baseline SPACE-domain data were available for 475,134 of 501,935 eligible participants (94.7%). Missingness across domains was low: Sleep, 1,076 (0.2%); Pain, 922 (0.2%); Affect, 9,199 (1.8%); Cognition, 4,964 (1.0%); and Energy, 17,239 (3.4%). Given the low proportion of missing domain data, complete-case analysis was used for primary structural analyses and multiple imputation was not undertaken.

Sex-stratified and age-restricted analyses were conducted as secondary analyses, including analyses in women aged ≤55 years to evaluate consistency in a subgroup relevant to perioperative and postpartum recovery research.

### 2.2 Ethics

UK Biobank has ethical approval from the North West Multi-centre Research Ethics Committee, and all participants provided written informed consent. This study was conducted under UK Biobank application number 791918 (Principal Investigator S.C.).

### 2.3 Sex, gender, race and ethnicity

Sex was defined using the UK Biobank sex variable (field 31), reflecting sex recorded at recruitment. Sex-and gender-based analyses were limited to sex-stratified models, and residual conflation of sex and gender cannot be excluded. Race and ethnicity were self-reported and treated as social constructs; ethnicity was collapsed to White versus non-White for analytic stability, this simplification limits inference about specific minoritised groups.

### 2.4 Conceptual framework

The analysis was guided by a framework (Figure S1) in which age, sex, socioeconomic deprivation, body mass index, smoking, comorbidity, early adversity, and polygenic liability were conceptualised as upstream factors that may influence both SPACE symptom burden and chronic pain or functional impairment. Work disability, self-rated health, and longstanding illness or disability were treated as concurrent clinical validation outcomes. Multimodal variables - actigraphy, biomarkers, imaging-derived phenotypes, and polygenic risk scores - were evaluated as external validation anchors.

### 2.5 Covariate selection and model adjustment

Covariates were selected a priori on the basis of their hypothesised associations with symptom burden and functional impairment. Primary adjusted models included age, sex, body mass index, Townsend Deprivation Index, and smoking status. Ethnicity was not included in the primary models because it was available only in a restricted subset; sensitivity analyses additionally adjusted for self-reported ethnicity, collapsed as White versus non-White, among participants with available data.

### 2.6 SPACE domain construction

SPACE domains were constructed a priori from baseline UK Biobank measures. Indicator selection was theory-informed and based on conceptual relevance to the SPACE framework, clinical interpretability, availability at baseline assessment, and suitability for population-level operationalisation.

Where multiple potential indicators were available within a domain, preference was given to measures reflecting broad, common, and clinically meaningful symptom burden rather than rare, context-specific, or exposure-dependent behaviours. General sleep-disturbance indicators were therefore prioritised over items such as falling asleep while driving. Low mood was selected as a primary affective indicator because it reflects clinically relevant affective burden and aligns with the conceptual aims of the framework.

All SPACE domains were derived from assessment-centre instance-0 variables. Component variables were oriented so that higher values indicated greater symptom burden, standardised as z-scores, and aggregated using row-wise means to generate domain-level scores.

The Sleep domain was derived from deviation from optimal sleep duration, defined as the absolute difference from 7 hours (field 1160); insomnia frequency (field 1200); and daytime dozing (field 1220). The Pain domain was derived from recent pain-site count (field 6159), with higher scores indicating broader pain burden across anatomical sites. The Affect domain incorporated neuroticism score (field 20127), happiness (field 4526), and frequency of depressed mood over the past two weeks (field 2050). The Cognition domain included fluid intelligence (field 20016), numeric memory (maximum digits remembered; field 4282), and mean reaction time for correct matches (field 20023). Cognitive variables were oriented so that higher values reflected greater impairment. Extreme values were winsorised before standardisation to reduce the influence of outliers. The Energy domain was based on self-reported tiredness or lethargy frequency over the past two weeks (field 2080).

Three composite SPACE scores were derived. SPACE-5 comprised all five domains. SPACE-4 comprised Sleep, Affect, Cognition, and Energy, omitting Pain. A four-domain SPAE score comprised Sleep, Pain, Affect, and Energy, excluding Cognition. SPAE was examined because preliminary analyses suggested the available UK Biobank cognition measures - primarily objective or performance-based - captured a partially distinct dimension. SPACE-4 tested whether the non-pain domains were associated with work disability independently of pain, given that work disability may itself arise from pain; SPACE-5 quantified the additional contribution of the Pain domain.

### 2.7 Structural evaluation of SPACE

The internal structure of SPACE was evaluated using correlation analysis, principal component analysis (PCA), exploratory factor analysis (EFA), and unsupervised clustering. Pearson correlations between standardised baseline domains were displayed as a heatmap. PCA was performed on standardised baseline domain scores among participants with complete data, reporting loadings and variance explained for all components. EFA used maximum-likelihood estimation: a one-factor model was fitted to SPACE-5 and a second to SPAE (excluding Cognition), with fit evaluated using the Tucker–Lewis Index (TLI), root mean square error of approximation (RMSEA), root mean square residual (RMSR), loadings, and variance explained, to assess whether Cognition behaved as part of the common factor.

### 2.8 SPACE phenotype clustering

Unsupervised k-means clustering used standardised baseline domain scores among participants with complete SPACE data. The number of clusters was selected using silhouette analysis. Cluster profiles were summarised using mean standardised domain scores and radar plots. Between-cluster differences were described using means with standard deviations or counts with percentages; given the large sample, standardised mean differences (SMDs) were prioritised over p values.

### 2.9 Clinical outcomes

The primary outcome was current work disability. Secondary outcomes included chronic pain interference, poor self-rated health, and longstanding illness or disability. Chronic pain interference was defined as pain interfering with usual activities during the preceding month and lasting for more than 3 months, thereby incorporating both chronicity and functional impact (Nicholas et al., 2019; Treede et al., 2019). It was defined using UK Biobank baseline pain fields. Participants first identified pain experienced during the preceding month that interfered with their usual activities (field 6159). For each reported pain type, the corresponding site-specific follow-up field was used to determine whether the pain had lasted for more than 3 months, including headache, facial pain, neck or shoulder pain, back pain, abdominal pain, hip pain, knee pain, and pain all over the body. A binary outcome was then derived: participants were classified as having chronic pain interference if at least one interfering pain type had persisted for more than 3 months, and as not having chronic pain interference if they selected “None of the above” or reported pain types for which all available duration responses were negative. Participants with “Prefer not to answer” responses or otherwise insufficient information were coded as missing.

Current work disability was defined from baseline employment status (field 6142) using the response “unable to work because of sickness or disability”; participants responding “none of the above” or “prefer not to answer” were excluded from this outcome analysis. Self-rated health status was derived from overall health rating (field 2178). Longstanding illness or disability was defined using self-reported longstanding illness, disability or infirmity (field 2188).

### 2.10 Exploratory multimodal evaluation

Secondary multimodal analyses in modality-specific subsets assessed convergent and incremental associations rather than causal mediation. Anchors included polygenic risk scores (PRS), structural brain MRI imaging-derived phenotypes, wrist actigraphy, and blood or physiological biomarkers. PRS were examined against continuous SPACE scores and high-burden phenotype membership, adjusted for age, sex, genotyping array, and ancestry principal components. Brain MRI imaging-derived phenotypes were analysed with adjustment for relevant technical and biological covariates, volumetric measures adjusted for intracranial volume where appropriate. An actigraphy composite was derived from mean overall acceleration, its standard deviation, and average moderate-to-vigorous physical activity (MVPA). Biomarkers were grouped into prespecified metabolic, inflammatory, and physiological indices, log-transformed where appropriate, standardised, and analysed against SPACE scores and phenotype membership, with false-discovery-rate correction.

### 2.11 Statistical analysis

Descriptive statistics summarised baseline characteristics overall and by phenotype, with SMDs for between-group magnitude. Associations of individual domains, composite scores, principal components, and phenotype membership with outcomes were examined using regression appropriate to outcome type - logistic regression was used for binary outcomes, including current work disability, poor self-rated health, longstanding illness or disability, pain interfering with usual activities in the preceding month and lasting for more than 3 months, and higher-burden phenotype membership. Sensitivity analyses additionally adjusted for ethnicity among participants with available ethnicity data. The incremental contribution of SPACE to each outcome was assessed primarily using likelihood-ratio tests comparing nested models (base model, base+SPACE-4, base+SPACE-5), which test whether the added terms contribute conditional information. Discrimination was summarised using the area under the receiver operating characteristic curve (AUC) with 95% confidence intervals, and the change in discrimination between nested models (ΔAUC) with bootstrap 95% confidence intervals, to convey the magnitude and precision of the increment rather than statistical significance alone in this very large sample; paired DeLong comparisons are reported secondarily. Because the analyses were intended to quantify concurrent associations and incremental discrimination rather than to develop an individual-level clinical prediction tool, calibration and clinical-utility analyses were not undertaken. The SPACE domains, their scoring, and the nested model sequence were prespecified and fixed before any analysis of these outcomes, and no data-driven predictor selection was performed; given the very large sample and small number of estimated parameters, overfitting was expected to be minimal. Sex-stratified and age-restricted analyses (including women aged ≤55 years) evaluated consistency. Secondary analyses examined consistency by age and sex. Sensitivity analyses additionally adjusted for ethnicity in the subset with available data. Analyses were performed in R (Posit/RStudio).

## 3 Results

### 3.1 Participant characteristics and completeness of SPACE-domain data

Among 501,935 eligible participants aged ≥40 years, complete baseline SPACE-domain data were available for 475,134 (94.7%) (participant flow, Figure 1). Missingness was low across all domains (Sleep 0.2%, Pain 0.2%, Affect 1.8%, Cognition 1.0%, Energy 3.4%). Primary structural analyses used complete cases.

**Figure 1.**
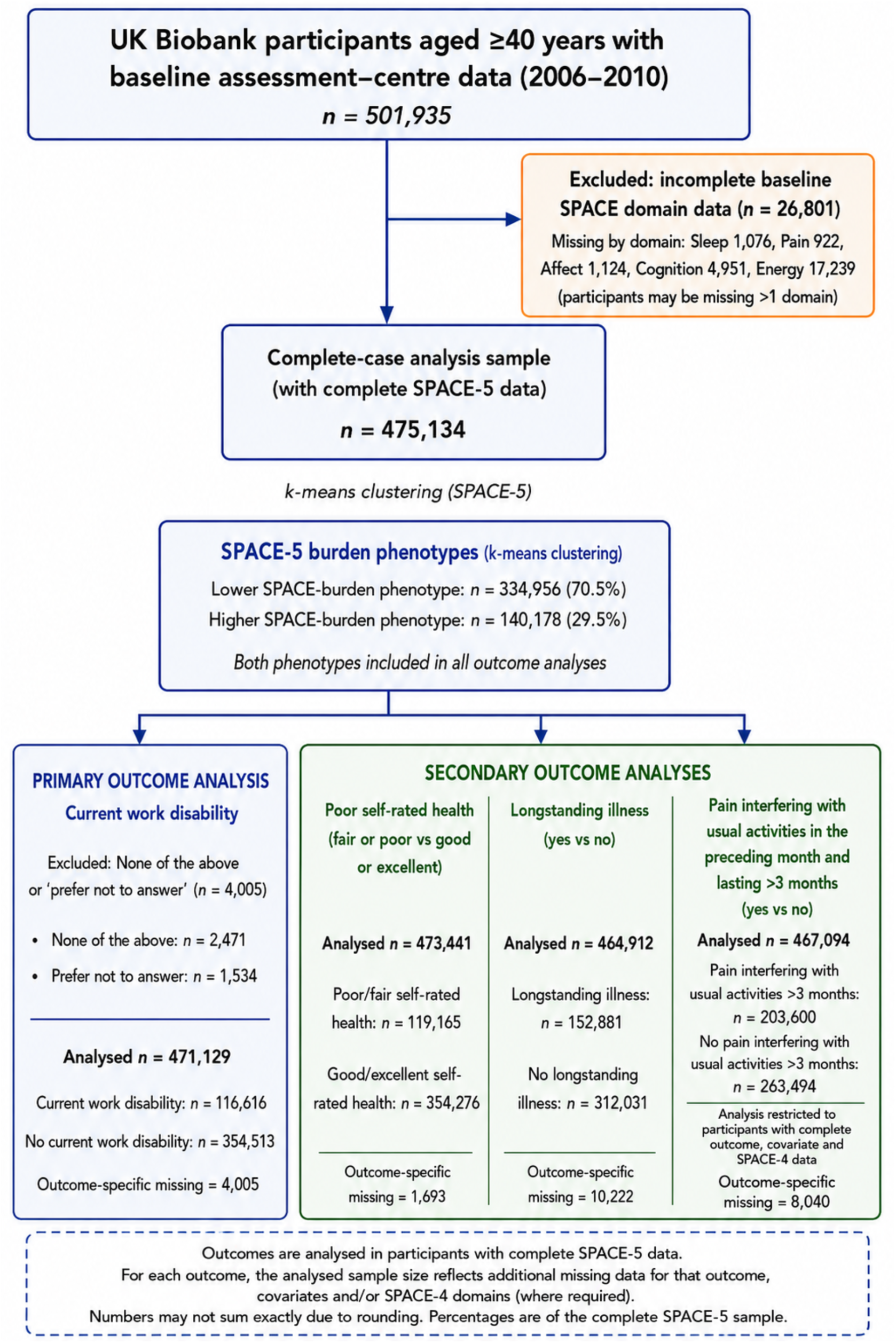
Participant flow and analytic samples. UK Biobank participants aged ≥40 years with baseline assessment-centre data were eligible for inclusion. Participants with incomplete baseline SPACE domain data were excluded from the complete-case SPACE-5 sample used for clustering and subsequent outcome analyses. K-means clustering identified lower- and higher-burden SPACE-5 phenotypes. Outcome-specific analytic samples varied because of missing outcome data and, where applicable, missing covariate or SPACE-4 data. Current work disability was the primary outcome; poor self-rated health, longstanding illness or disability, and chronic pain interference were secondary outcomes. Chronic pain interference was defined as pain interfering with usual activities during the preceding month and lasting for more than 3 months. Numbers may not sum exactly because participants could have missing data for more than one variable. SPACE, Sleep, Pain, Affect, Cognition and Energy.

3.2 SPACE phenotype clustering

Silhouette analysis supported a two-cluster solution distinguishing lower- and higher-burden phenotypes (Table 1; Figure 2&3). The lower-burden cluster included 334,956 participants (70.5%) and the higher-burden cluster 140,178 (29.5%). The higher-burden phenotype showed elevated mean standardised scores across all domains, particularly Energy (0.97), Affect (0.82), Pain (0.54), and Sleep (0.45), with a smaller elevation for Cognition (0.14); the lower-burden cluster showed lower scores throughout. Symptom variation was thus organised primarily along a lower-to-higher burden gradient rather than into discrete subtypes.

**Figure 2.**
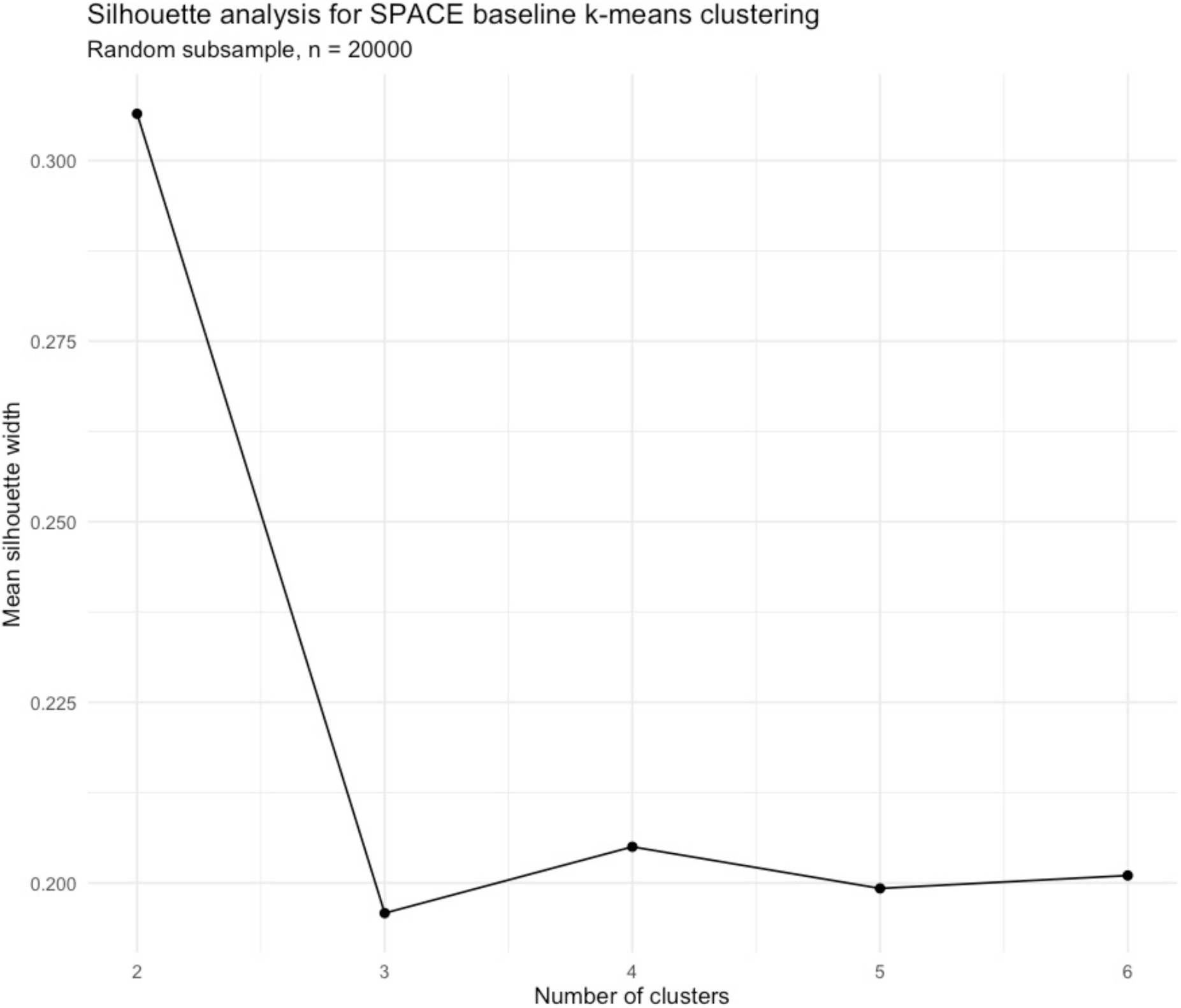
Silhouette analysis for selection of the number of SPACE burden clusters. Mean silhouette width was calculated for k-means solutions containing two to six clusters in a random subsample of 20,000 participants with complete baseline SPACE-5 data. The two-cluster solution had the highest mean silhouette width and was therefore selected for the primary clustering analysis. SPACE, Sleep, Pain, Affect, Cognition and Energy.

**Table 1.** UK Biobank SPACE k-means cluster profiles (mean standardised domain scores) Mean standardised scores for the Sleep, Pain, Affect, Cognition and Energy domains are shown for the two clusters identified by k-means analysis among participants with complete baseline SPACE-5 data. Higher values indicate greater domain burden. SPACE, Sleep, Pain, Affect, Cognition and Energy.

| Cluster | n | % | Sleep | Pain | Affect | Cognition | Energy |
| --- | --- | --- | --- | --- | --- | --- | --- |
| Lower SPACE burden | 334,956 | 70.5 | −0.19 | −0.48 | −0.32 | −0.09 | −0.42 |
| Higher SPACE burden | 140,178 | 29.5 | 0.45 | 0.54 | 0.82 | 0.14 | 0.97 |

### 3.3 Baseline characteristics by SPACE phenotype

Compared with the lower-burden phenotype, the higher-burden phenotype was more likely to be female (61.8% vs 51.1%) and had higher body mass index (28.36 vs 27.02), greater deprivation (Townsend -0.71 vs -1.62), more current smoking (14.2% vs 8.9%), and less regular physical activity (48.6% vs 57.0%); mean age was similar (Table 2). The largest between-group differences were for functional variables: current work disability (10.3% vs 1.3%), disability living allowance or blue-badge status (12.0% vs 2.4%), and longstanding illness or disability (48.8% vs 24.6%). Given the sample size, p values were uniformly small and less informative than SMDs, which were largest for these functional variables, deprivation, and body mass index.

**Table 2.**
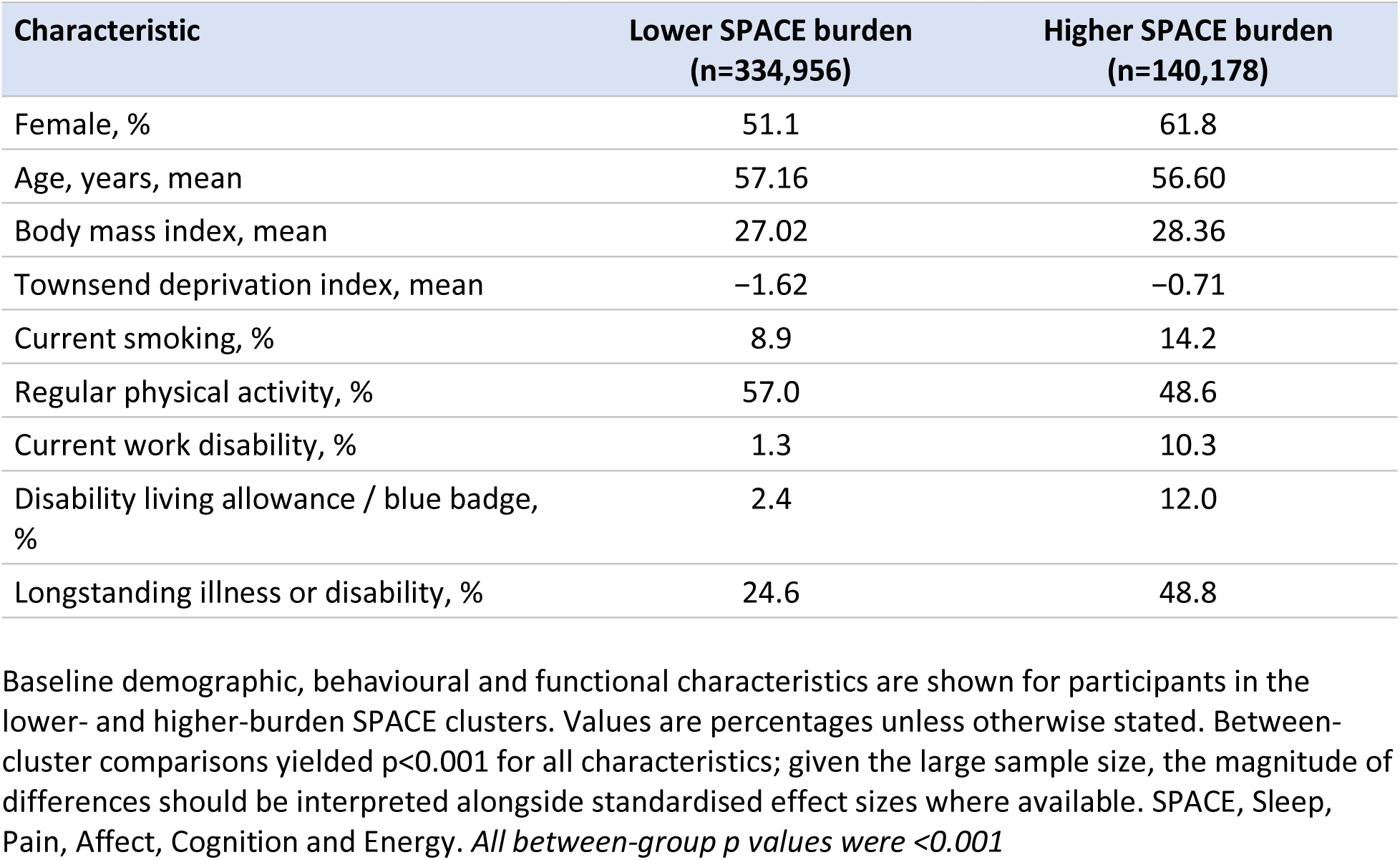
Baseline characteristics by SPACE burden cluster. Baseline demographic, behavioural and functional characteristics are shown for participants in the lower- and higher-burden SPACE clusters. Values are percentages unless otherwise stated. Between-cluster comparisons yielded p<0.001 for all characteristics; given the large sample size, the magnitude of differences should be interpreted alongside standardised effect sizes where available. SPACE, Sleep, Pain, Affect, Cognition and Energy. *All between-group p values were <0.001*

### 3.4 Correlation structure of SPACE domains

Baseline domains showed positive but generally modest correlations (r=0.04-0.46; Figure 4), consistent with a multidimensional symptom-system rather than interchangeable indicators. The strongest association was between Affect and Energy. Cognition showed weaker correlations with the other domains, indicating partial independence within the broader structure.

**Figure 3.**
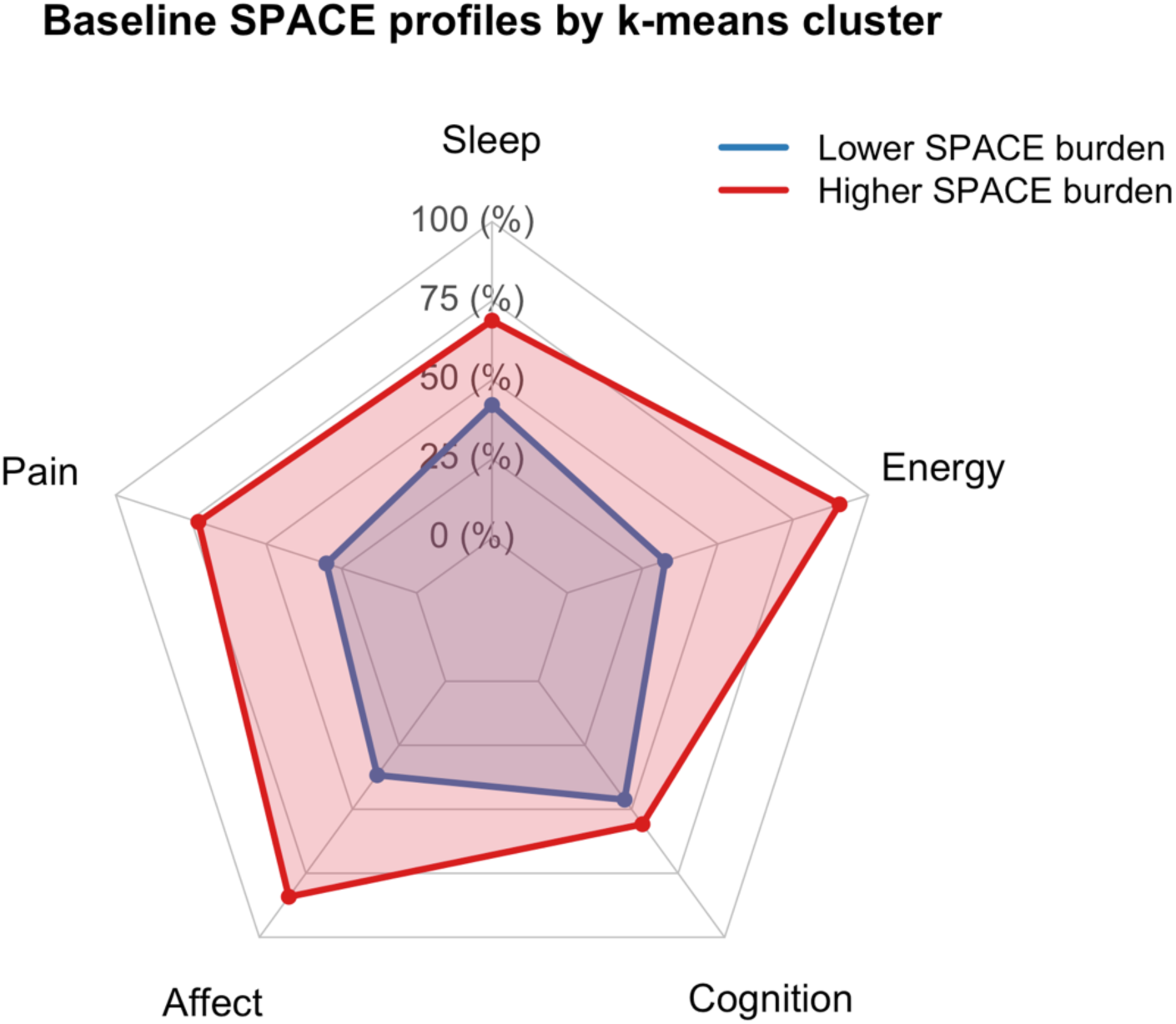
Baseline SPACE domain profiles by k-means cluster. Radar plot showing the relative burden across Sleep, Pain, Affect, Cognition and Energy domains in the lower- and higher-burden SPACE clusters. Domain values were rescaled to percentages for visualisation, with higher values indicating greater symptom burden. The higher-burden cluster showed consistently greater burden across all five domains, with the largest separation for Energy and Affect and the smallest separation for Cognition. SPACE, Sleep, Pain, Affect, Cognition and Energy.

**Figure 4.**
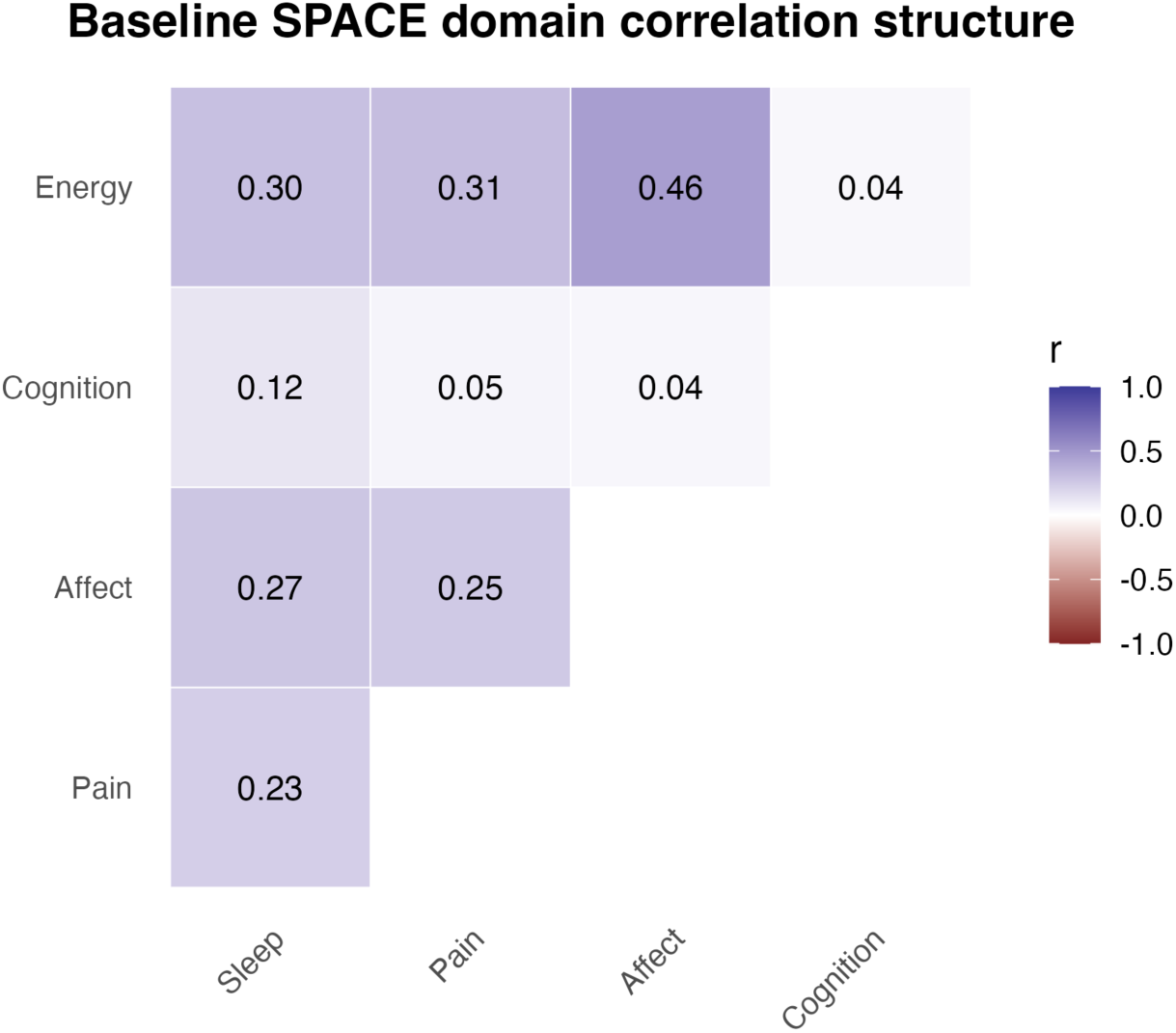
Correlation structure of baseline SPACE domains. Heatmap showing pairwise Pearson correlations between standardised Sleep, Pain, Affect, Cognition and Energy domain scores among participants with complete baseline SPACE-5 data. Correlations were positive but generally modest, with the strongest association observed between Affect and Energy (r=0.46). Cognition showed comparatively weak correlations with the other domains. SPACE, Sleep, Pain, Affect, Cognition and Energy.

### 3.5 Principal component analysis of SPACE domains

The first principal component (PC1) explained 38.8% of variance with positive loadings across all five domains, consistent with a shared symptom-burden dimension; Energy, Affect, Sleep, and Pain contributed most strongly (0.554, 0.528, 0.451, 0.442), whereas Cognition loaded weakly (0.128) (Table 3). PC2 explained 20.2% and was dominated by Cognition (0.943). The first two components together explained 59.0% of variance. SPACE thus contained both a shared symptom-burden dimension and domain-specific structure, with the available cognition measures capturing a partially distinct dimension.

**Table 3.** Principal component analysis of baseline SPACE domains. Loadings of the five standardised SPACE domains on principal components 1–5 are shown, together with eigenvalues, percentage variance explained and cumulative variance explained. Larger absolute loadings indicate a stronger contribution of a domain to the corresponding component. The sign of a loading indicates direction and is arbitrary with respect to component orientation. SPACE, Sleep, Pain, Affect, Cognition and Energy.

| Domain | PC1 | PC2 | PC3 | PC4 | PC5 |
| --- | --- | --- | --- | --- | --- |
| Sleep | 0.451 | 0.214 | -0.121 | -0.857 | -0.046 |
| Pain | 0.442 | -0.069 | 0.880 | 0.098 | -0.127 |
| Affect | 0.528 | -0.168 | -0.408 | 0.328 | -0.647 |
| Cognition | 0.128 | 0.943 | -0.020 | 0.304 | 0.026 |
| Energy | 0.554 | -0.176 | -0.211 | 0.237 | 0.750 |
| **Eigenvalue** | 1.938 | 1.010 | 0.776 | 0.746 | 0.531 |
| **Variance explained** | 38.8% | 20.2% | 15.5% | 14.9% | 10.6% |
| **Cumulative variance** | 38.8% | 59.0% | 74.5% | 89.4% | 100.0% |

### 3.6 Exploratory factor analysis of SPACE domains

In the one-factor SPACE-5 model, Energy and Affect loaded most strongly (0.716, 0.632), Sleep and Pain moderately (0.435, 0.434), and Cognition weakly (0.087); fit was acceptable (TLI 0.942; RMSEA 0.055; RMSR 0.033) and the model explained 25.9% of variance. The SPAE model (excluding Cognition) again showed strongest loadings for Energy and Affect (0.721, 0.631) and moderate loadings for Sleep and Pain (0.430, 0.432), with improved fit (TLI 0.973; RMSEA 0.047; RMSR 0.021) and 32.2% of variance explained. These findings support Sleep, Pain, Affect, and Energy as an empirically cohesive core, with Cognition retained on conceptual grounds. The weaker empirical integration of Cognition likely reflects the use of objective, performance-based measures (reaction time, fluid intelligence, numeric memory) rather than subjective cognitive dysfunction.

### 3.7 Associations with and discrimination of current work disability

Higher burden in each SPACE domain was associated with greater odds of current work disability across age and sex strata (Table 4). In SPACE-4 models, Sleep showed the strongest association in participants aged <55 years (OR per 1-SD increase 2.67, 95% CI 2.58–2.77) and those aged ≥55 years (2.07, 2.01–2.13). Affect, Cognition, and Energy were also independently associated with work disability in both age groups. Associations were similar in women and men. In SPACE-5 models, Pain was independently associated with work disability, while associations with the four non-pain domains were retained. All domain associations had P<0.001.

**Table 4.** Associations between SPACE domains and current work disability, stratified by age and sex By age. Odds ratios are from multivariable logistic regression and represent the change in odds of current work disability per 1-SD increase in the respective domain. SPACE-4 includes Sleep, Affect, Cognition, and Energy; SPACE-5 additionally includes Pain, measured using recent pain-site count. Domains were entered simultaneously within each model. Age-stratified models were adjusted for continuous age, sex, body mass index, Townsend deprivation index, and current smoking. Sex-stratified models were adjusted for age, body mass index, Townsend deprivation index, and current smoking. In a sensitivity analysis restricted to women with available ethnicity data, additional adjustment for ethnicity produced materially unchanged estimates. OR, odds ratio; CI, confidence interval; SD, standard deviation.

| Domain | Age <55 y, OR<br>(95% CI) | P | Age ≥55 y, OR<br>(95% CI) | P |
| --- | --- | --- | --- | --- |
| **SPACE-4** |  |  |  |  |
| Sleep | 2.67 (2.58–2.77) | <0.001 | 2.07 (2.01–2.13) | <0.001 |
| Affect | 1.44 (1.40–1.47) | <0.001 | 1.29 (1.26–1.32) | <0.001 |
| Cognition | 1.36 (1.32–1.39) | <0.001 | 1.29 (1.26–1.31) | <0.001 |
| Energy | 1.42 (1.39–1.46) | <0.001 | 1.51 (1.48–1.54) | <0.001 |
| **SPACE-5** |  |  |  |  |
| Sleep | 2.55 (2.45–2.64) | <0.001 | 1.94 (1.88–2.00) | <0.001 |
| Pain | 1.37 (1.34–1.40) | <0.001 | 1.53 (1.50–1.56) | <0.001 |
| Affect | 1.38 (1.35–1.42) | <0.001 | 1.22 (1.20–1.25) | <0.001 |

| Domain | Age <55 y, OR (95% CI) | P | Age ≥55 y, OR (95% CI) | P |
| --- | --- | --- | --- | --- |
| Cognition | 1.36 (1.32–1.40) | <0.001 | 1.29 (1.26–1.32) | <0.001 |
| Energy | 1.35 (1.32–1.39) | <0.001 | 1.40 (1.37–1.43) | <0.001 |

By sex
| Domain | Female, OR (95% CI) | P | Male, OR (95% CI) | P |
| --- | --- | --- | --- | --- |
| **SPACE-4** |  |  |  |  |
| Sleep | 2.39 (2.31–2.47) | <0.001 | 2.09 (2.02–2.15) | <0.001 |
| Affect | 1.35 (1.32–1.38) | <0.001 | 1.39 (1.36–1.42) | <0.001 |
| Cognition | 1.27 (1.24–1.30) | <0.001 | 1.30 (1.27–1.33) | <0.001 |
| Energy | 1.50 (1.47–1.54) | <0.001 | 1.45 (1.42–1.48) | <0.001 |
| **SPACE-5** |  |  |  |  |
| Sleep | 2.27 (2.19–2.34) | <0.001 | 1.97 (1.90–2.03) | <0.001 |
| Pain | 1.44 (1.41–1.46) | <0.001 | 1.48 (1.45–1.51) | <0.001 |
| Affect | 1.29 (1.26–1.32) | <0.001 | 1.33 (1.30–1.36) | <0.001 |
| Cognition | 1.27 (1.24–1.30) | <0.001 | 1.31 (1.28–1.34) | <0.001 |
| Energy | 1.41 (1.38–1.44) | <0.001 | 1.36 (1.33–1.38) | <0.001 |

**Table 5.** Model discrimination (AUC with 95% CI) for clinical validation outcomes. Base model: included age, sex, body mass index, Townsend deprivation index and smoking. **Pain-interference outcome:** was evaluated using SPACE-4 only because including the Pain domain in SPACE-5 would introduce direct criterion overlap with the outcome. **Ethnicity sensitivity analysis:** was performed in the subset with ethnicity available in the current export. Adding ethnicity produced negligible changes in AUC: work disability, +0.000004 (base), -0.000052 (SPACE-4) and -0.000023 (SPACE-5); poor self-rated health, +0.00369, +0.000893 and +0.000641, respectively; longstanding illness, +0.000068, +0.000013 and +0.000057, respectively. **Statistical comparisons:** The incremental contribution of SPACE-4 and SPACE-5 over the base model was significant for every non-overlapping outcome by likelihood-ratio test (P<0.001); paired DeLong comparisons were reported secondarily. **Abbreviations:** AUC, area under the receiver operating characteristic curve; CI, confidence interval.

| Outcome | Base model AUC (95% CI) | SPACE-4 AUC (95% CI) | SPACE-5 AUC (95% CI) |
| --- | --- | --- | --- |
| Current work disability | 0.748 [0.745, 0.752] | 0.850 [0.847, 0.853] | 0.863 [0.860, 0.866] |
| Current poor self-rated health | 0.688 [0.686, 0.690] | 0.788 [0.787, 0.790] | 0.804 [0.802, 0.805] |
| Current longstanding illness | 0.645 [0.643, 0.646] | 0.698 [0.696, 0.699] | 0.718 [0.716, 0.719] |
| Pain interfering with usual activities in the preceding month and lasting >3 months | 0.593 [0.591, 0.595] | 0.672 [0.671, 0.674] | Not evaluated |

At the model level, adding SPACE improved discrimination of current work disability beyond the five-covariate base model, from an AUC of 0.748 (95% CI 0.745–0.752) to 0.850 (0.847–0.853) with SPACE-4 and 0.863 (0.860–0.866) with SPACE-5; the incremental contribution was supported by likelihood-ratio tests (both P<0.001).

### 3.8 Chronic pain interference and other secondary outcomes

The same pattern held for poor self-rated health (0.688→0.788→0.805) and longstanding illness (0.645→0.698→0.718), with the non-pain SPACE-4 domains contributing most of the gain. Among 467,094 participants included in the complete-case analysis, 203,600 (43.6%) had at least one pain type that had interfered with usual activities during the preceding month and had lasted for more than 3 months. In the adjusted SPACE-4 model, the strongest associations were observed for Energy (OR per 1-SD increase 1.42, 95% CI 1.41–1.43) and Sleep (OR 1.40, 95% CI 1.38–1.41), followed by Affect (OR 1.24, 95% CI 1.23–1.25); Cognition showed only a small association (OR 1.01, 95% CI 1.00–1.02; P=0.010). Addition of the four SPACE domains improved model fit compared with the five-covariate model (likelihood-ratio χ²[4]=30,415, P<0.001). The five-covariate model showed limited discrimination (AUC 0.593, 95% CI 0.591–0.595), while addition of SPACE-4 increased the AUC to 0.672 (95% CI 0.671–0.674), an absolute increase of 0.079 (paired DeLong P<0.001). SPACE-5 was not evaluated because including the Pain domain would introduce direct criterion overlap with the outcome.

### 3.9 Exploratory multimodal associations

Convergence with objective measures was assessed in women aged ≤55 years using wrist actigraphy, biomarkers, PRS, and MRI-derived phenotypes; this subgroup was selected for relevance to perioperative and postpartum recovery research and does not imply the framework is specific to women (Ciechanowicz and Ma, 2025; Ciechanowicz et al., 2026). A reduced demographic base model discriminated current longstanding illness, disability, or infirmity poorly (AUC 0.575). Adding SPACE substantially improved discrimination (AUC 0.678; ΔAUC +0.103). Sequential addition of actigraphy (0.691), MRI-derived phenotypes (0.695), PRS (0.700), and metabolic and inflammatory composites (0.709) each added modest information, so symptoms provided the principal discriminatory contribution (Supplementary Tables S1-S3).

Higher SPACE burden aligned with reduced or altered physical-activity profiles on actigraphy and with adverse metabolic and inflammatory markers, including triglycerides (OR 1.15), waist-to-hip ratio (1.14), body-fat percentage (1.14), glycated haemoglobin (1.10), white blood cell count (1.08), neutrophil count (1.07), C-reactive protein (1.06), lymphocyte count (1.05), and neutrophil-to-lymphocyte ratio (1.03); composite metabolic (OR 1.22) and inflammatory (1.09) indices were also associated with high-burden membership. Genetic analyses showed associations with polygenic susceptibility for schizophrenia (OR 1.07/SD), bipolar disorder (1.05), and age at menopause (0.95). Exploratory brain MRI analyses identified associations with thalamic volume, cortical grey-matter volume, bilateral cortical volume, frontal medial cortical grey-matter volume, and right amygdala measures. Given the exploratory design and modest incremental value, these results are hypothesis-generating and require confirmation with full effect-size reporting.

## 4 Discussion

This population-scale analysis indicates that chronic pain interference and functional impairment occur within a broader context of multidimensional symptom burden across Sleep, Pain, Affect, Cognition, and Energy. SPACE showed evidence of a shared symptom-burden dimension while retaining domain-specific variation. Domain-based modelling and unsupervised clustering identified a higher-burden subgroup with markedly worse functional outcomes, including a substantially higher prevalence of current work disability. These findings support interpretation of SPACE as a clinically interpretable multidimensional symptom profile rather than a set of interchangeable symptoms or a single measure of distress.

The association with chronic pain interference is particularly relevant. Addition of the four non-pain domains improved discrimination of pain that had interfered with usual activities and persisted for more than 3 months, despite exclusion of the Pain domain to avoid direct criterion overlap. Sleep, Energy, and Affect showed the strongest associations, whereas Cognition contributed only modestly. This suggests that the functional impact of chronic pain may be embedded within a broader context incorporating disturbed sleep, fatigue, and affective burden. It is consistent with the original identification of SPACE as a symptom-severity dimension distinct from generalised sensory sensitivity in chronic overlapping pain conditions (Schrepf et al., 2018). It also complements longitudinal UK Biobank evidence that a greater number of central nervous system-associated symptoms predicts subsequent chronic primary pain (Kelleher et al., 2025b). The present cross-sectional results cannot establish that non-pain symptoms cause pain interference, but indicate that pain-related limitation is not fully characterised by pain location or intensity alone.

The magnitude of association with current work disability was also notable. Participants in the higher-burden phenotype had substantially more work disability than those in the lower-burden phenotype, and each domain remained independently associated with work disability when entered simultaneously. Sleep showed the strongest association, particularly among participants younger than 55 years. Sleep disturbance may affect occupational functioning through daytime alertness, emotional regulation, physical endurance, and cognitive performance. However, the findings must not be interpreted directionally. Work-limiting illness may worsen sleep, fatigue, mood, and perceived function, just as high multidomain symptom burden may make continued employment more difficult. Both may also reflect underlying disease severity or socioeconomic adversity.

Our findings complement previous chronic-pain phenotyping studies. Hsu and colleagues identified UK Biobank chronic-pain subgroups based on age, sex, and number of pain sites, reporting shared problems involving sleep, mobility, and daily functioning, while fatigue and depression varied particularly across widespread-pain phenotypes (Hsu et al., 2025). The present study is conceptually distinct because it adopts a symptom-first rather than pain-location-first approach. Multidomain symptom burden identified a higher-impact phenotype associated with both chronic pain interference and disability, suggesting that clinically important pain morbidity may reflect broader vulnerability architectures not captured by pain site count alone.

The structural analyses further refine interpretation of SPACE. Correlations between domains were positive but generally modest, while the first principal component captured a common symptom-burden dimension. Exploratory factor analysis supported a relatively cohesive core comprising Sleep, Pain, Affect, and Energy. Cognition was less strongly integrated and loaded predominantly on a separate principal component. This does not necessarily mean that cognition is clinically irrelevant. The UK Biobank indicators represented objective performance, including reaction time, fluid intelligence, and numeric memory, whereas the cognition component of symptom frameworks is generally closer to subjective cognitive difficulty or perceived executive function. The weaker loading may therefore reflect measurement mismatch. Future studies should compare subjective and performance-based cognitive measures directly.

These results also caution against treating SPACE solely as a unidimensional score. Although the analyses identified a shared burden gradient, the domains were not redundant and their relative associations differed between outcomes. Sleep and Energy were prominent for chronic pain interference and work disability, while Pain added information for work disability. Cognition was structurally distinct and contributed minimally to chronic pain interference in the available operationalisation. An overall score may therefore identify high cumulative burden, whereas domain-level profiles retain clinical interpretability and may reveal different targets for intervention.

The multimodal findings were directionally concordant but modest. Higher SPACE burden was associated with less favourable physical-activity, metabolic, inflammatory, genetic, and brain-imaging characteristics, but these measures added relatively little discrimination beyond the symptom domains. One interpretation is that self-reported symptoms integrate the accumulated functional consequences of multiple biological, psychological, and social processes more effectively than any single objective marker. Alternatively, the limited increments may reflect restricted modality-specific samples, measurement error, selected markers, or the exploratory design.

Previous UK Biobank work reported sex-specific associations between chronic pain and imaging-derived phenotypes, including cortical and striatal measures (Parisien et al., 2025). In the present study, women in the higher-burden phenotype showed differences in thalamic and cortical volume measures, with additional exploratory associations in regions implicated in affective and default-mode processing.

These findings provide preliminary evidence that multidomain symptom burden may align with distributed biological variation. Similarly, associations with metabolic and inflammatory indices were generally small and may reflect shared upstream determinants, including adiposity, physical inactivity, smoking, deprivation, and comorbidity. The multimodal findings should therefore be interpreted as hypothesis-generating.

The findings may also be relevant to perioperative and recovery research. Studies of chronic postsurgical pain after caesarean delivery have traditionally focused on obstetric, perioperative, and pain-related factors, including acute postoperative pain and analgesic requirements (Sng et al., 2009; Jin et al., 2016; Borges et al., 2020). Preliminary work applying SPACE in the postpartum setting has suggested that multidomain symptom burden may be associated with persistent pain after caesarean delivery (Ciechanowicz and Ma, 2025; Ciechanowicz et al., 2026). The present findings do not establish translation to postpartum or surgical populations, but support the premise that persistent morbidity may arise within a multidimensional symptom state. This interpretation is compatible with longitudinal postpartum studies showing that sleep, physical activity, sedentary behaviour, and recovery-related symptoms evolve together (Evenson et al., 2012; Horwitz et al., 2023; Jessen et al., 2025; Pandal et al., 2025). Prospective validation in perioperative and postpartum cohorts is required before SPACE can be used for risk stratification or clinical decision-making.

The clinical implication is not that SPACE should replace pain assessment, diagnostic evaluation, or condition-specific measures. Rather, assessment of functionally impairing pain may benefit from extending beyond pain intensity, duration, and anatomical distribution. Sleep disturbance, fatigue, affective burden, and cognitive symptoms may identify additional dimensions of impairment relevant to daily and occupational function. A multidimensional symptom profile could therefore complement conventional pain assessment by characterising the wider context in which pain becomes disabling.

Future research should determine whether baseline SPACE burden predicts incident chronic pain interference, work loss, healthcare use, or impaired recovery; whether changes in SPACE domains correspond to changes in function; and whether particular profiles are associated with treatment response. External validation is required in more diverse populations and in cohorts using purpose-built measures. Prospective analyses are also needed to distinguish whether SPACE acts as a risk marker, consequence of disability, or concurrent representation of underlying multisystem burden.

### 4.1 Strengths and limitations

Strengths include the large, well-characterised cohort, prespecified domain construction, complementary structural analyses, clinically meaningful outcomes, and exploratory evaluation across actigraphy, biomarkers, genetics, and brain imaging. Exclusion of the Pain domain from the chronic pain-interference analysis reduced direct criterion overlap and enabled evaluation of whether non-pain symptoms were associated with pain-related impairment.

Several limitations warrant emphasis. First, the main exposure and outcomes were assessed cross-sectionally, precluding causal and temporal inference. The relationship between symptom burden and disability is plausibly bidirectional, and both may reflect shared underlying disease or social determinants.

Second, the SPACE domains and clinical outcomes were largely self-reported at the same assessment and covered partially overlapping aspects of health. Some discrimination may therefore reflect shared-method variance or construct overlap, particularly for self-rated health and longstanding illness or disability. Chronic pain interference incorporates functional limitation by definition, although omission of the Pain domain reduced direct exposure–outcome overlap.

Third, increases in area under the curve should not be interpreted as evidence of clinical utility. They quantify concurrent incremental discrimination within the same cohort. The prespecified domains and model sequence reduce the risk of data-driven optimisation but do not eliminate these limitations.

Fourth, pragmatic UK Biobank proxies were used rather than purpose-built SPACE instruments. This was especially important for Cognition, which was represented by performance-based testing rather than subjective cognitive symptoms. Fifth, multimodal analyses were exploratory, undertaken in modality-specific subsets, and partly restricted to women aged 55 years or younger. Finally, healthy-volunteer selection in UK Biobank may limit generalisability and may underestimate associations in populations with greater symptom and socioeconomic burden.

## 5 Conclusions

SPACE was identifiable as a multidimensional symptom profile at population scale and was concurrently associated with chronic pain interference and work disability. Functionally impairing pain occurred within a broader symptom context involving sleep disturbance, affective burden and low energy, rather than pain burden alone. Objective multimodal measures showed modest convergence but added relatively little discrimination beyond symptoms. Prospective external validation is required to determine whether SPACE predicts incident disability, impaired recovery, healthcare use or treatment response, and whether purpose-built domain measures improve its structural and clinical validity.

## Authorship contributions

Contributions are described using CRediT roles. S.C: Conceptualization; Methodology; Software; Formal analysis; Writing - original draft. K. L: Formal analysis; Writing - review & editing. D.M: Supervision; Writing - review & editing.

## Data Availability

Individual-level data are available from UK Biobank (https://www.ukbiobank.ac.uk) to approved researchers.

## Acknowledgements

Funding

European Society of Anesthesiology and Intensive Care (ESAIC_GR_2022_DM), Brussels, Belgium, and Ningbo Top Medical and Health Research Program (2024010317), Ningbo, China (to DMA).

## Study pre-registration

The analysis was conducted under an approved UK Biobank research application that prospectively specified investigation of biopsychosocial factors and health-related outcomes. However, the present analysis and complete statistical analysis plan were not formally preregistered. The SPACE domain structure was defined independently of the outcome analyses.

## Open materials

Derived variable definitions and analysis code needed to reproduce the reported analyses are available upon request.

## Reporting

The study is reported in accordance with STROBE (cross-sectional).

## Declarations

During the preparation of this work the author(s) used ChatGPT (OpenAI) and Claude (Anthropic) to assist with language editing and code review. All analyses and manuscript content were reviewed by the authors, who take full responsibility for the work.

## Supplementary Materials

**STROBE checklist: cross-sectional study**

| Item | Recommendation | Reported | Location / comment |
| --- | --- | --- | --- |
| 1a | Study design indicated in title or abstract | Yes | Both state the design: Title (“a cross-sectional... validation”) and Abstract (“In this cross-sectional analysis of UK Biobank...”). |
| 1b | Balanced abstract summary of what was done and found | Yes | Abstract (p.1–2). |
| 2 | Scientific background and rationale | Yes | Introduction (p.2–3). |
| 3 | Specific objectives / hypotheses | Yes | Introduction, final paragraph (p.3). |
| 4 | Key elements of study design early in the paper | Yes | Methods states cross-sectional design and same-visit ascertainment. |
| 5 | Setting, locations, relevant dates | Yes | Methods; UK Biobank baseline recruitment 2006–2010 stated. |
| 6a | Eligibility criteria; sources and selection of participants | Yes | Methods, Study design (p.3). |
| 7 | Outcomes, exposures, predictors, confounders, effect modifiers | Yes | Methods, SPACE construction / Outcomes / Covariates (p.4–6). |
| 8 | Data sources and measurement for each variable | Yes | Methods (field IDs) (p.5–6). |
| 9 | Efforts to address potential sources of bias | Yes | Covariate adjustment; shared-method/criterion-overlap and healthy-volunteer bias addressed in Methods and Limitations. |
| 10 | Study size | Yes | Analytic sample = maximal complete-case UK Biobank sample; derivation of numbers in Results and Figure 1. No formal sample-size calculation applies to this fixed resource. |
| 11 | Quantitative variables and groupings | Yes | z-standardisation, winsorising, composites (p.5); cluster groups (p.6). |
| 12a | Statistical methods, including confounding control | Yes | Methods, Statistical analysis (p.9). |
| 12b | Methods for subgroups and interactions | Yes | Sex- and age-stratified analyses (p.9). |
| 12c | How missing data were addressed | Yes | Complete-case; missingness quantified (p.3, p.9). |
| 12d | (Cross-sectional) sampling strategy accounting | N/A | Single-wave baseline sample; not a complex survey design. |
| 12e | Sensitivity analyses | Yes | Sensitivity analyses described (p.9). |
| 13a | Numbers of individuals at each stage | Yes | Results (501,931 → 475,134) (p.9). |
| 13b | Reasons for non-participation | Yes | Exclusions (incomplete SPACE data, per-domain missingness) shown in Figure 1 and Results. |
| 13c | Flow diagram | Yes | Figure 1 (participant flow). |
| 14a | Characteristics of participants; exposures/confounders | Yes | Table 1; Results (p.13). |
| 14b | Number of participants with missing data per variable | Yes | Methods/Results (p.9). |
| 15 | Outcome events or summary measures | Yes | Outcome frequencies by phenotype (p.13). |
| 16a | Adjusted estimates with confidence intervals; confounders | Yes | Table 4 (p.13–14). |
| 16b | Category boundaries when continuous variables categorised | Yes | Cluster/phenotype definitions (p.6). |
| 16c | Translate relative to absolute risk where relevant | Yes | Absolute risks by phenotype reported (Table 1; e.g., work disability 1.3% vs 10.3%). |
| 17 | Other analyses (subgroups, interactions, sensitivity) | Yes | Multimodal and stratified analyses (p.14). |
| 18 | Key results with reference to objectives | Yes | Discussion (p.14). |
| 19 | Limitations | Yes | Discussion, Strengths and limitations (p.16). |
| 20 | Cautious overall interpretation | Yes | Discussion (p.14–16). |
| 21 | Generalisability | Yes | Healthy-volunteer bias discussed (p.16). |
| 22 | Funding and role of funders | Yes | Disclosures (p.16–17). |

---

**Supplementary Figure S1.**
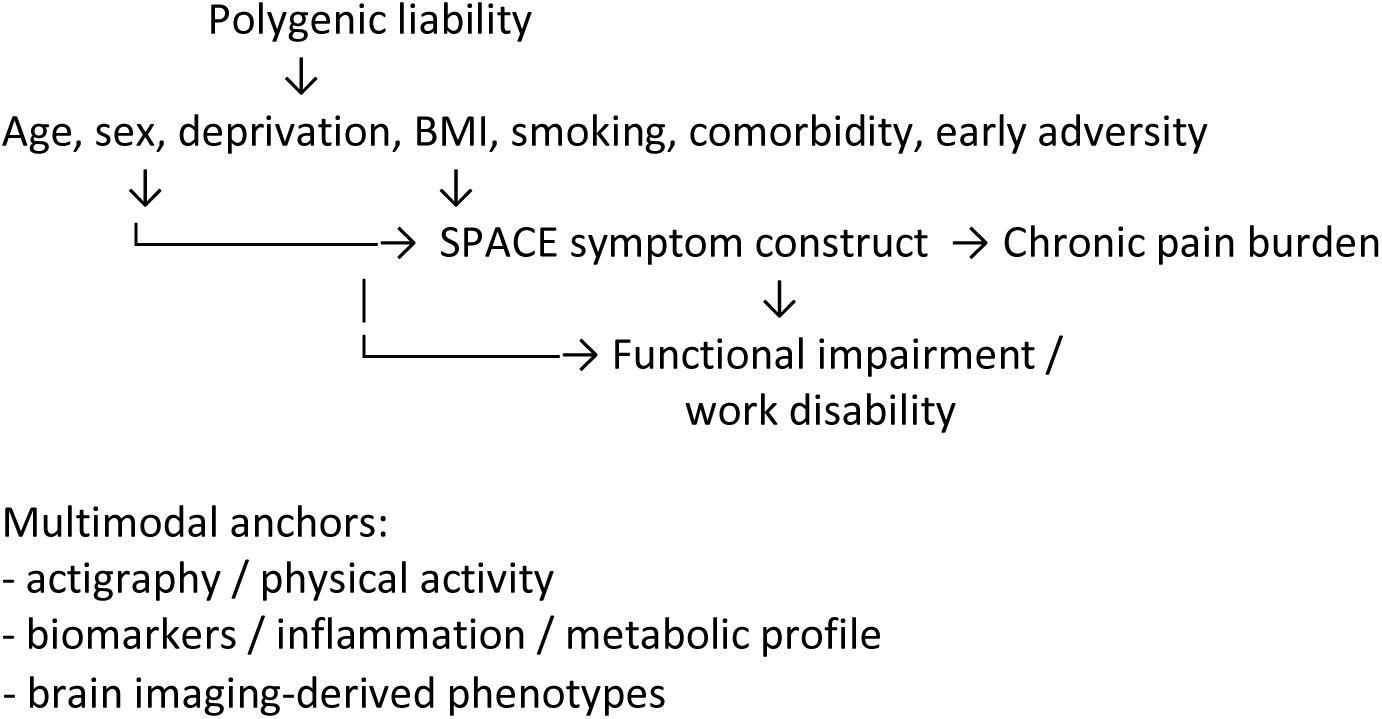
Conceptual framework for the hypothesised relationships guiding the analysis. Upstream factors (age, sex, deprivation, body mass index, smoking, comorbidity, early adversity, and polygenic liability) may influence both the SPACE symptom construct and the outcomes; SPACE (Sleep, Pain, Affect, Cognition, Energy) relates to chronic pain burden and, directly and via pain, to functional impairment/work disability. Multimodal measures (actigraphy/physical activity, inflammatory and metabolic biomarkers, brain imaging-derived phenotypes) are shown as external multimodal anchors. Arrows denote assumed, not established, directions of effect; the cross-sectional design precludes causal interpretation.

**Table S1.** Polygenic risk score (PRS) associations with high-burden SPACE phenotype

| PRS | OR per 1 SD | 95% CI | P value | FDR (BH) | n |
| --- | --- | --- | --- | --- | --- |
| Standard PRS: Schizophrenia (SCZ) | 1.075 | 1.054 to 1.096 | <0.0001 | 6.40e-12 | 55,935 |
| Standard PRS: Bipolar disorder (BD) | 1.060 | 1.039 to 1.081 | <0.0001 | 3.17e-08 | 55,935 |
| Enhanced PRS: Age at menopause (AAM) | 0.957 | 0.926 to 0.989 | <0.05 | 2.34e-02 | 18,785 |
| Enhanced PRS: Bipolar disorder (BD) | 1.045 | 1.011 to 1.080 | <0.05 | 2.34e-02 | 18,785 |
| Enhanced PRS: Schizophrenia (SCZ) | 1.046 | 1.012 to 1.082 | <0.05 | 2.34e-02 | 18,785 |
| Standard PRS: Age at menopause (AAM) | 0.980 | 0.961 to 0.999 | <0.05 | 6.86e-02 | 55,935 |
| Enhanced PRS: Body mass index (BMI) | 1.038 | 1.002 to 1.075 | <0.05 | 6.86e-02 | 18,785 |
| Standard PRS: Body mass index (BMI) | 0.981 | 0.961 to 1.001 | >=0.05 | 8.75e-02 | 55,935 |
| Standard PRS: Systemic lupus erythematosus (SLE) | 1.018 | 0.999 to 1.038 | >=0.05 | 9.31e-02 | 55,935 |
| Enhanced PRS: Systemic lupus erythematosus (SLE) | 1.028 | 0.995 to 1.063 | >=0.05 | 1.20e-01 | 18,785 |
| Standard PRS: Rheumatoid arthritis (RA) | 0.993 | 0.974 to 1.012 | >=0.05 | 4.73e-01 | 55,935 |
| Enhanced PRS: Rheumatoid arthritis (RA) | 1.012 | 0.979 to 1.047 | >=0.05 | 4.73e-01 | 18,785 |

Analytic cohort: females aged ≤55 years with PRS data: n = 60,413. High-burden n = 15,047; Logistic regression models estimated odds ratios (OR) for high-burden SPACE status per 1 SD increase in PRS, adjusted for age, BMI, Townsend, and PRS genetic principal components (p26201\_a0–a3). FDR is Benjamini–Hochberg across all PRS tested.

**Table S2.**
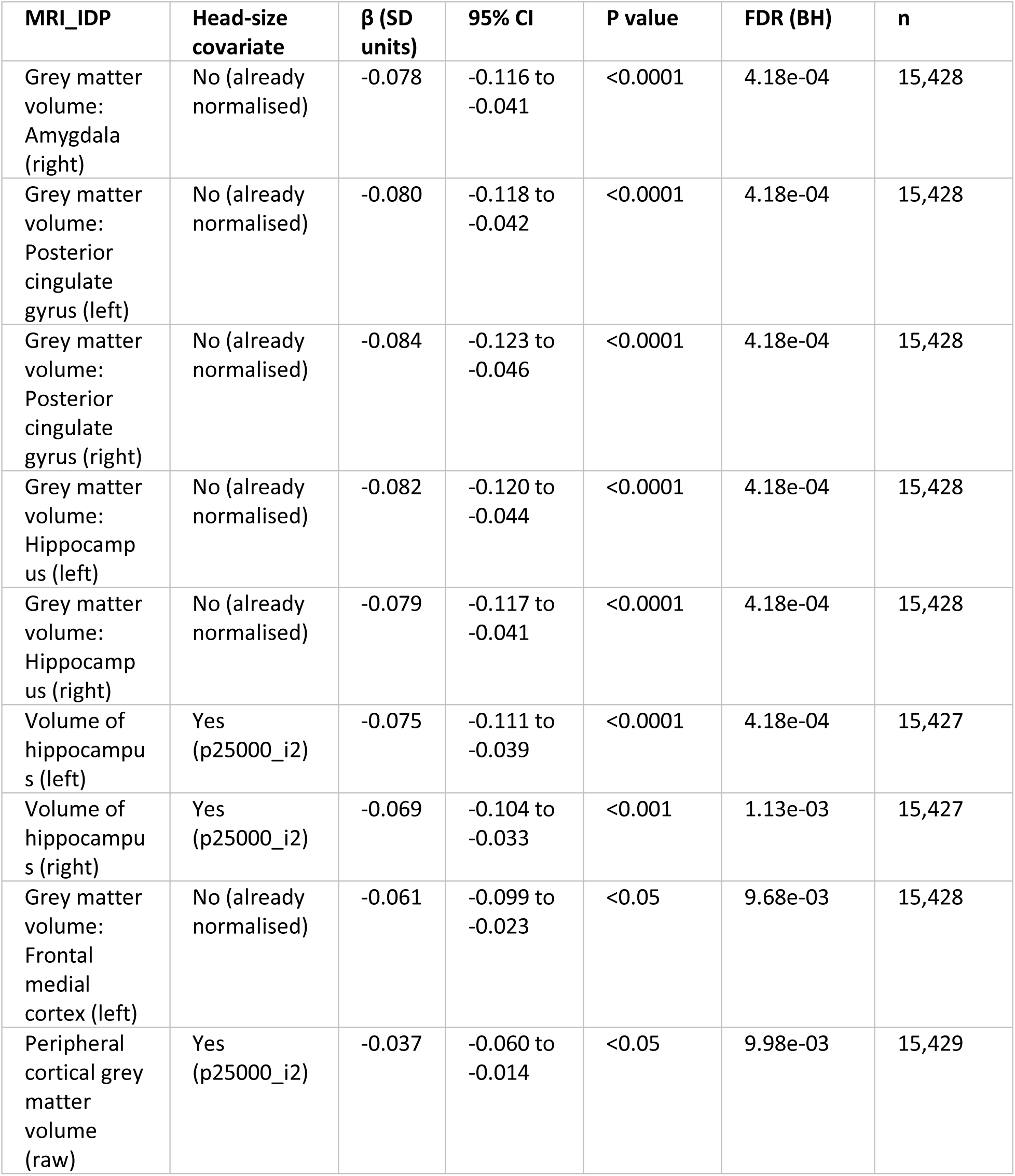
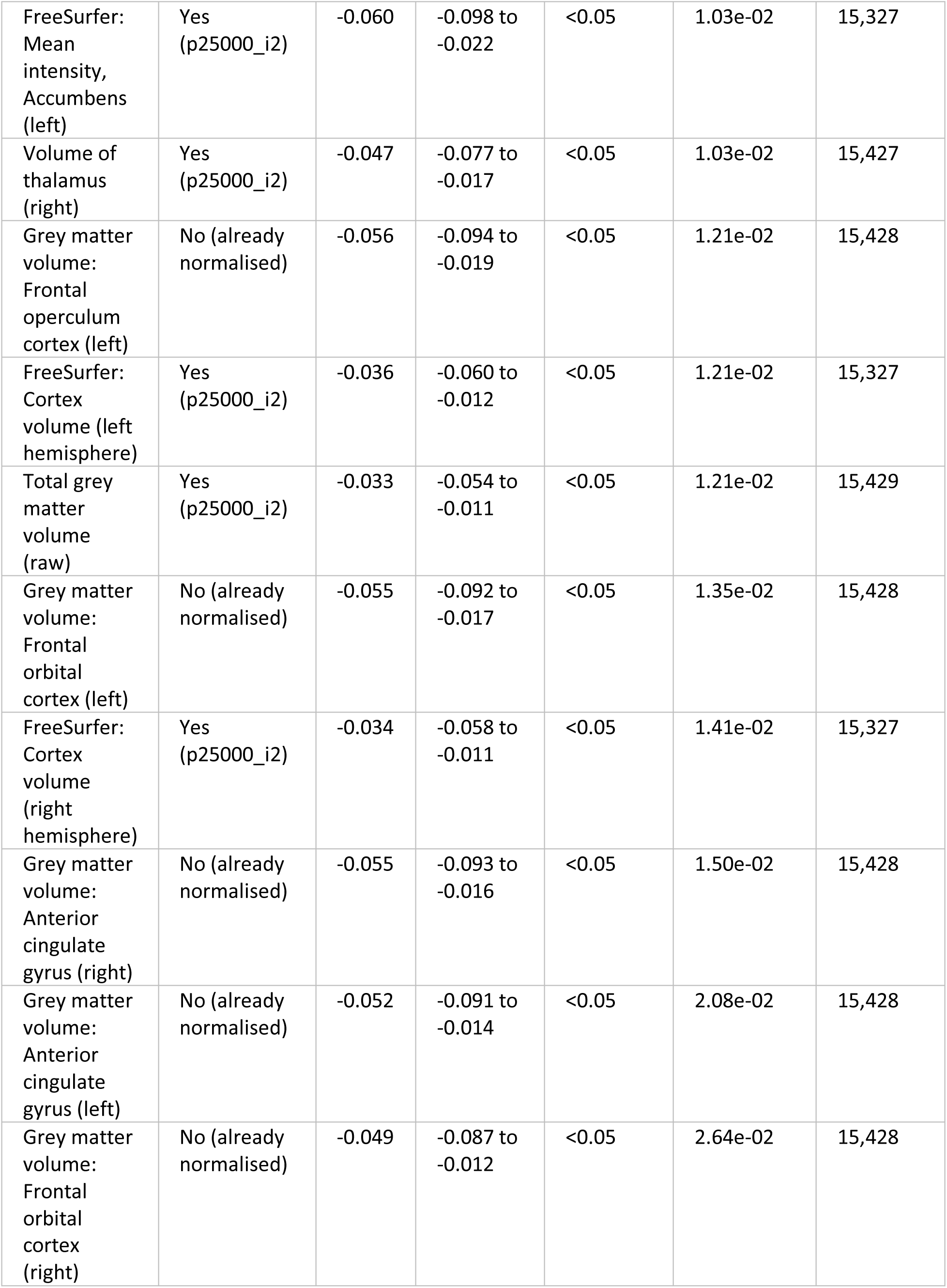
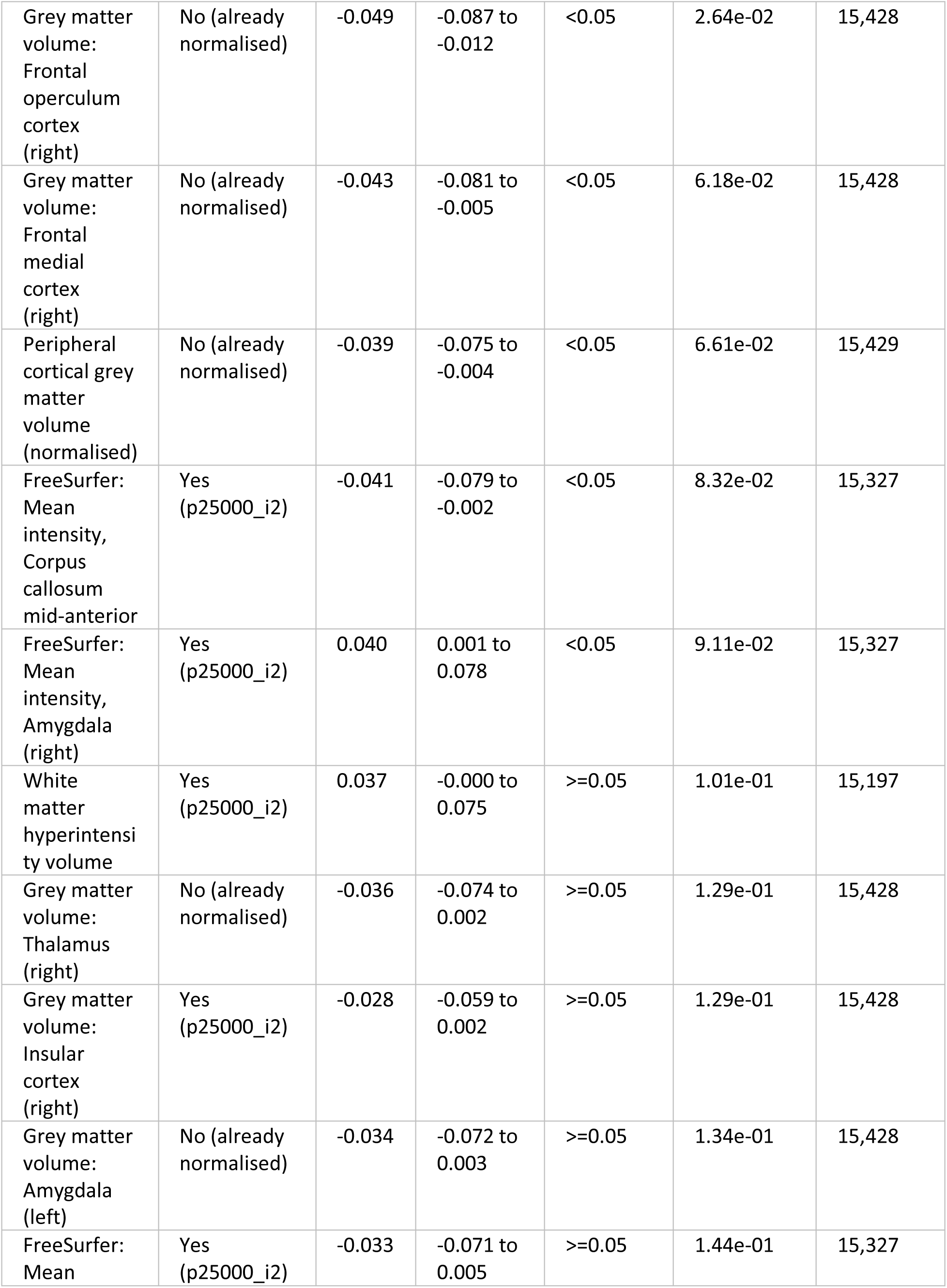
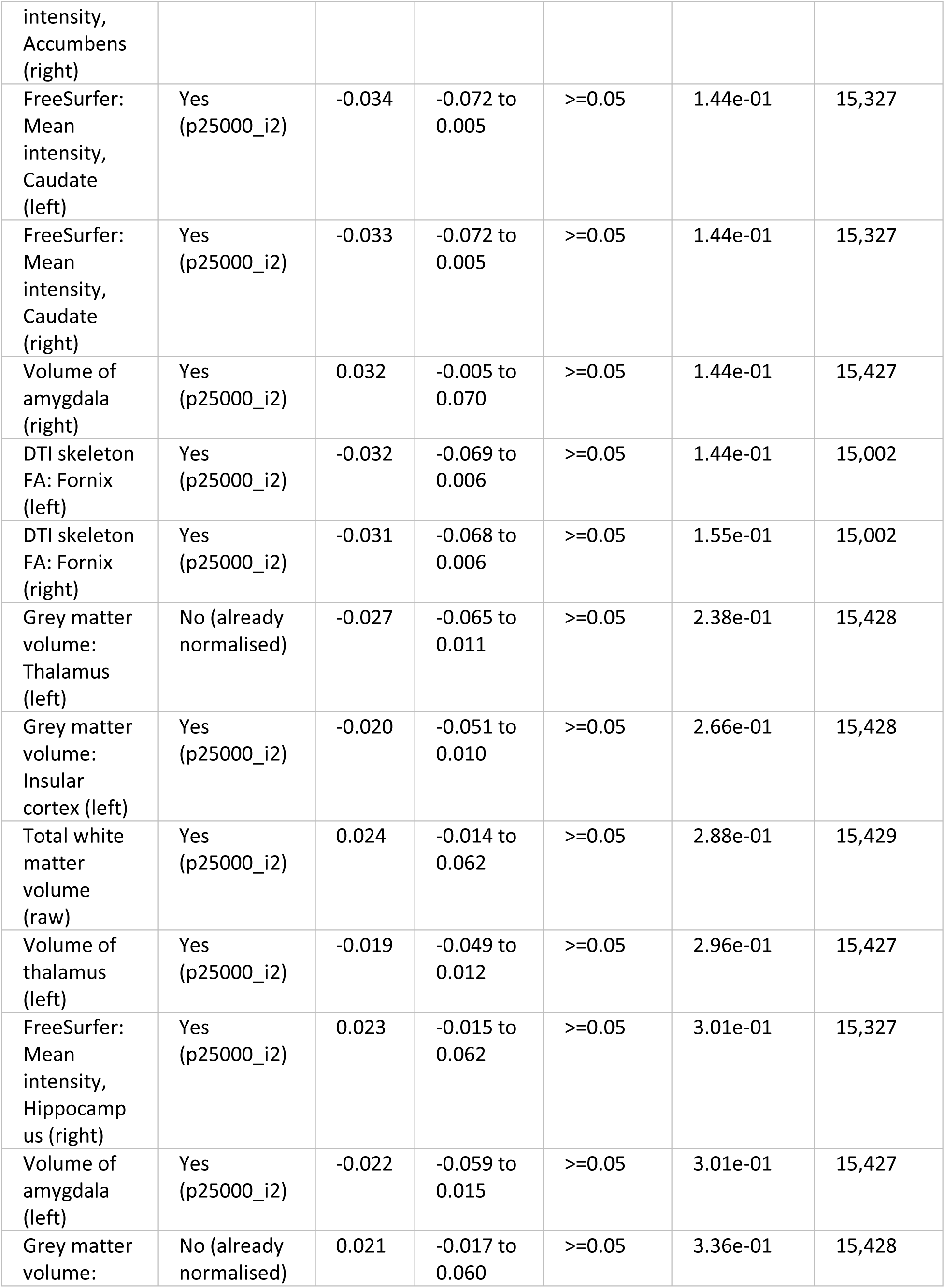
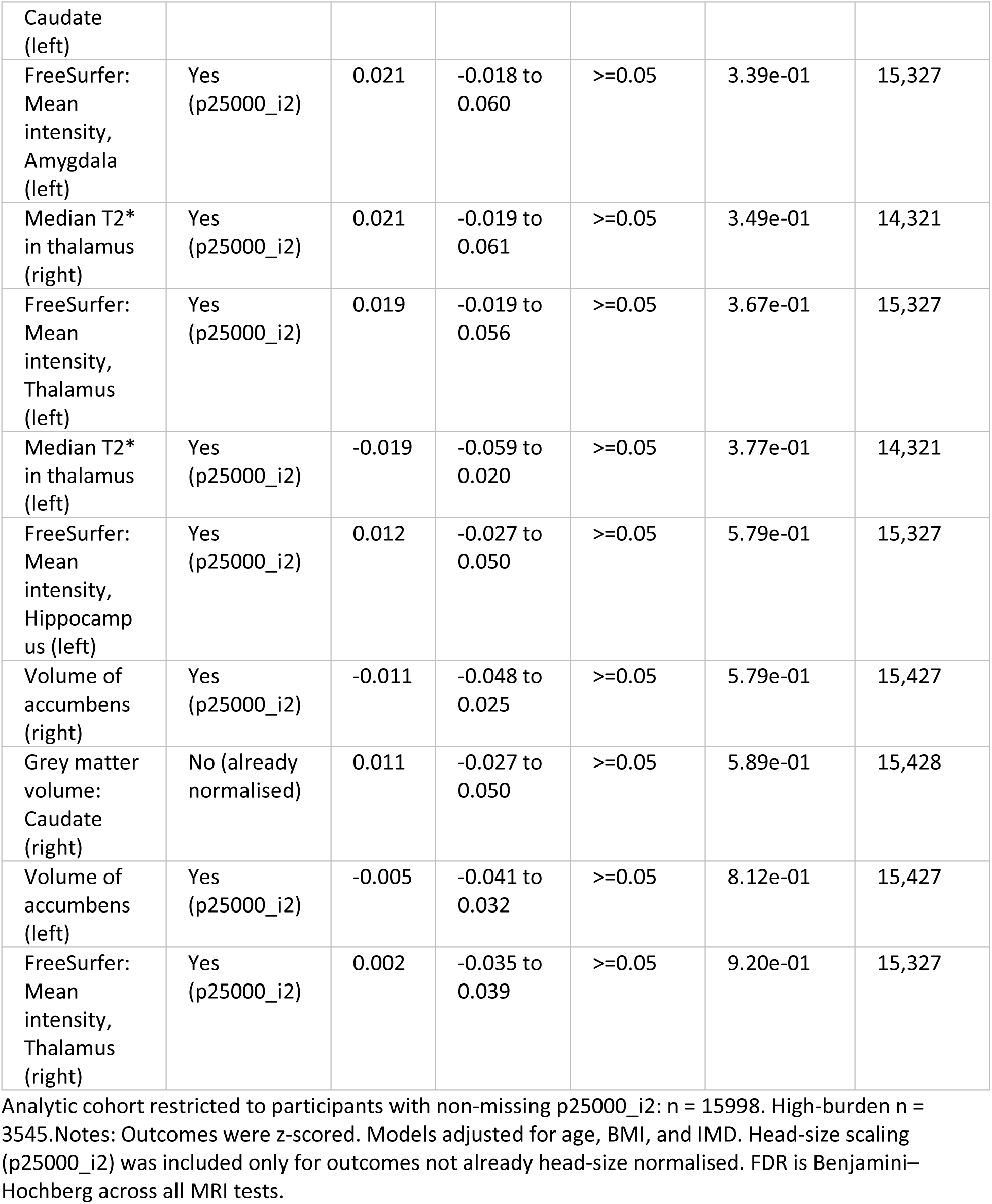
Associations between high-burden SPACE phenotype and MRI-derived phenotypes (UK Biobank imaging subsample of females aged 55 and under).

**Table S3.** Association Between High-Burden SPACE Phenotype and Biological Markers (females ≤55 years).

| Outcome | $\beta$ (95% CI) | P-value | N |
| --- | --- | --- | --- |
| **Metabolic composite (SD)** | 0.17 (0.16 to 0.18) | $p < 0.0001$ | 54,654 |
| HbA1c (mmol/mol) | 0.72 (0.62 to 0.82) | $p < 0.0001$ | 54,275 |
| log(Triglycerides) | 0.11 (0.10 to 0.12) | $p < 0.0001$ | 54,851 |
| LDL cholesterol (mmol/L) | 0.05 (0.04 to 0.07) | $p < 0.0001$ | 54,805 |
| HDL cholesterol (mmol/L) | -0.06 (-0.07 to -0.05) | $p < 0.0001$ | 49,511 |
| **Inflammatory composite (SD)** | 0.12 (0.10 to 0.13) | $p < 0.0001$ | 55,816 |
| log(CRP) | 0.22 (0.20 to 0.24) | $p < 0.0001$ | 54,800 |
| **Autonomic composite (SD)** | -0.02 (-0.04 to -0.00) | $p < 0.05$ | 32,994 |
| Systolic BP (mmHg) | -1.34 (-1.65 to -1.03) | $p < 0.0001$ | 55,263 |
| Arterial stiffness index | 0.14 (0.08 to 0.20) | $p < 0.001$ | 33,250 |
| Resting heart rate (bpm) | 2.45 (1.49 to 3.41) | $p < 0.001$ | 2,569 |

Values represent adjusted $\beta$ coefficients for high-burden SPACE cluster membership. The metabolic composite comprised waist-to-hip ratio, body mass index (BMI), body fat percentage, glycated haemoglobin (HbA1c), log-transformed triglycerides, LDL cholesterol, total cholesterol, and HDL cholesterol (reverse coded). The inflammatory composite comprised log-transformed C-reactive protein (CRP), total white cell count, and log-transformed neutrophil-to-lymphocyte ratio. The autonomic composite comprised systolic blood pressure, diastolic blood pressure, arterial stiffness index, and resting heart rate (Winsorised at the 1st and 99th percentiles prior to standardisation). Composite outcomes are expressed in standard deviation units. Metabolic and inflammatory models were adjusted for age and Index of Multiple Deprivation (IMD). Autonomic models were adjusted for age, BMI, and IMD.

